# COVID-19 trends and severity among symptomatic children aged 0-17 years in ten EU countries, 3 August 2020 – 3 October 2021

**DOI:** 10.1101/2021.11.25.21266875

**Authors:** Nick Bundle, Nishi Dave, Anastasia Pharris, Gianfranco Spiteri, Charlotte Deogan, Jonathan E. Suk, study group members

## Abstract

To guide evidence-based prevention of COVID-19 in children, we estimated risks of severe outcomes in 820,404 symptomatic paediatric cases reported by 10 EU Member States between August 2020 and October 2021. Case and hospitalisation rates rose as overall transmission increased but severe outcomes were rare: 9,611 (1.2%) were hospitalised, 640 (0.08%) required intensive care and 84 (0.01%) died. Despite increased individual risk (aOR; 95% CI for hospitalisation: 7.3; 3.3 - 16.2, ICU: 8.7; 6.2 - 12.3) in cases with comorbidities such as cancer, diabetes, cardiac or lung disease, most (83.7%) hospitalised children had no reported comorbidity.

## Manuscript

### Understanding the burden of COVID-19 in children in Europe

Understanding the burden of COVID-19 among children is essential for evidence-based decision-making regarding the vaccination of children, as well as for assessing the importance of SARS-CoV-2 mitigation measures in specific settings, such as in schools [1]. Here, we report on the burden and severity of symptomatic notified COVID-19 cases among children in the EU.

We analysed, using R v4.1.1, pooled case-based surveillance data reported to the European Surveillance System (TESSy) by 10 EU countries (Austria, Cyprus, Finland, Germany, Ireland, Italy, Luxembourg, Malta, Slovakia and Sweden) for symptomatic COVID-19 cases (reported as symptomatic or with a date of onset), between weeks 32-2020 and 43-2021. Data were restricted to weeks 32-2020 to 39-2021 to account for delayed reporting of the outcomes: hospitalisation, ICU (admission to ICU and/or requiring ventilation or ECMO) or death. We compared the cumulative number of reported deaths by each country [2] to official data from public sources [3], with a minimum 90% completeness threshold for study inclusion. Time series of reported hospital and ICU admissions were compared to those of admission or occupancy data obtained from public sources[4], excluding countries with reported time series that were incomplete or whose peaks occurred at different times. Outcomes reported as ‘unknown’ were recoded to ‘no’ for deaths from all countries, and for hospitalisation/ICU from Ireland (following national practice) and Sweden (all cases were coded as ‘yes’ or ‘unknown’). Hospitalisation/ICU status was recoded to ‘no’ if date of hospitalisation preceded date of onset to minimise inclusion of incidental hospital admissions in the crude risk numerators. Remaining cases with unknown outcome or sex were excluded.

We described trends in pooled weekly rates and proportions by age group of notified symptomatic cases and hospital admissions for the entire study period. Age-specific cumulative case notification rates per 100 000 population and crude risk for hospitalisation, ICU (among all cases and among hospitalised cases) and death were estimated by age group in years and in months <2 years using a subset of data from seven countries (Austria, Cyprus, Finland, Ireland, Luxembourg, Slovakia and Sweden) reporting age in months. We considered the overall period and the period in which the delta variant of concern was dominant, based on country-specific estimates of when alpha-delta co-circulation ceased (defined as alpha falling to 5% of reported weekly sequences, median weeks 28-2021 to 39-2021). Age-specific distributions of the outcomes between period and by sex within the overall period were compared using Chi-squared tests.

A subset of data from seven countries (Cyprus, Finland, Italy, Luxembourg, Malta, Slovakia and Sweden) reporting data on comorbidities (coded in TESSy as cancer, diabetes, cardiac disease, lung disease, neuromuscular disease; HIV, asthma, kidney disease, hypertension, pregnancy, liver disease, obesity; smoker/history of smoking) was analysed to estimate AR stratified by presence or absence of any comorbidities, having removed cases with unknown comorbidity. We used logistic regression to estimate age-specific (<1, 1-4, 5-11, 12-17) and overall (0-17 years) adjusted odds ratios (aOR) for each outcome for cases with and without any comorbidity.

### Trends in case notification and hospitalisation

Weekly notification rates among symptomatic cases increased sharply in all age groups in EU/EEA countries from July 2021 (Figure 1a). Hospitalisation rates also increased in all age groups in line with cases, but from much lower levels in children than in adults (Figure 1c). Since January 2021, children have represented an increasing proportion of total reported cases and hospital admissions although the proportion of hospitalisations have stabilised somewhat since July (Figure 1b, d).

**Figure 1:**
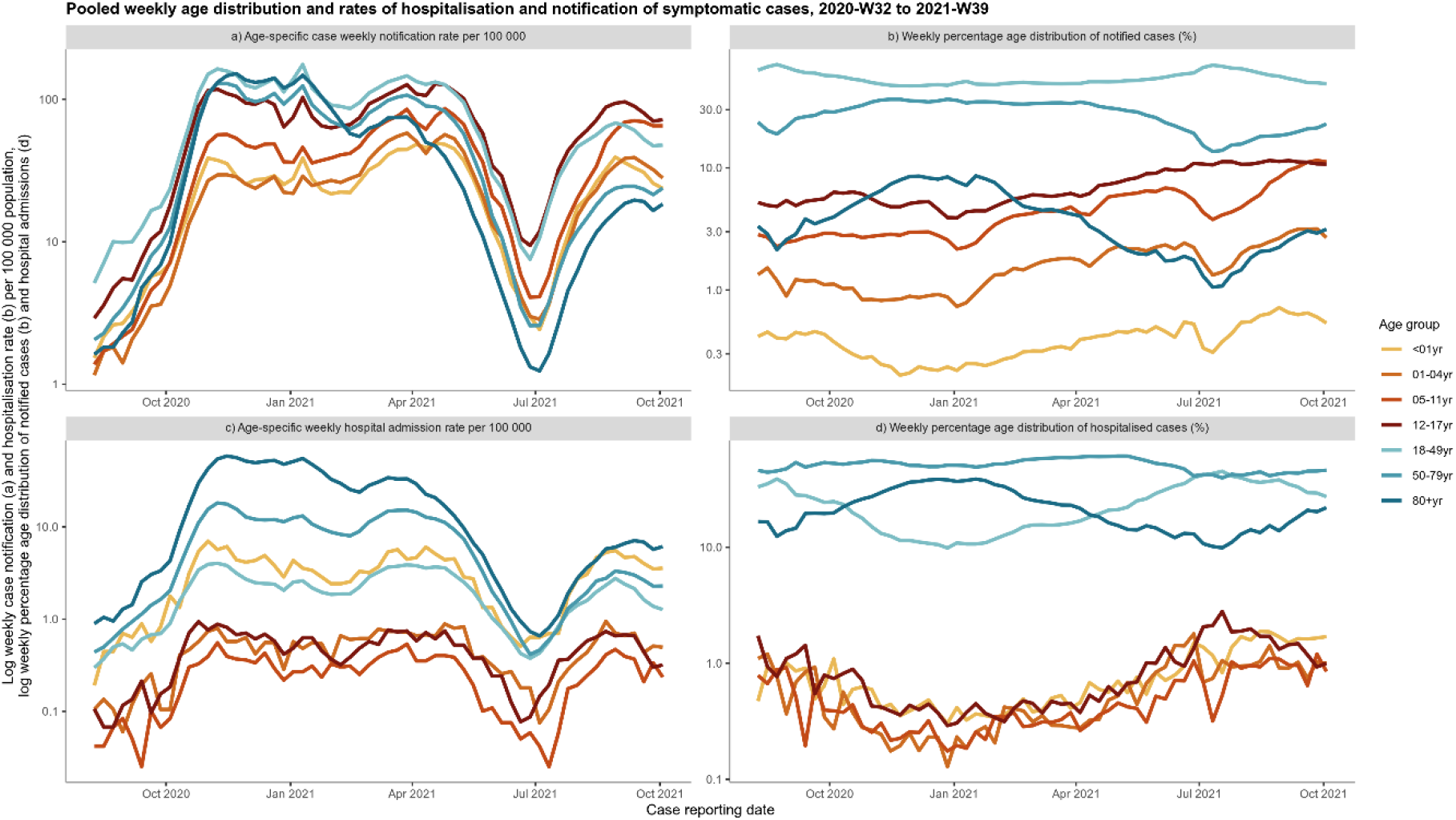
Weekly age distribution and rates of notified symptomatic COVID-19 cases and hospital admissions reported to TESSy, pooled from 10 EU countries, weeks 32-2020 to 39-2021.

During the period 32-2020 to 39-2021, cumulative case rates reported among a total of 820,404 symptomatic paediatric cases (12.4% of 6,604,483 cases of all ages) increased with every additional year of age from 2-17 years (Table 1). Hospitalisation was reported for 9,611 (1.2%) cases, ICU for 640 (0.08% of all cases, 6.7% of hospitalised cases) and death for 84 (0.01%).

**Table 1:** Age-specific counts and crude risks of severe COVID-19 outcomes by age, pooled from 10 EU countries, weeks 32-2020 to 39-2021.

| Age | Symptomatic Cases | Cases per 100k population | Hospitalisation |  | ICU |  | Death |  | ICU among hospitalised cases |
| --- | --- | --- | --- | --- | --- | --- | --- | --- | --- |
|  |  |  | n | Crude risk % (95% CI) | n | Crude risk % (95% CI) | n | Crude risk % (95% CI) | Crude risk % (95% CI) |
| **Total 0-23 months (1)** | **8258** |  | **695** | **8.42 (7.83 - 9.04)** | **95** | **1.15 (0.93 - 1.40)** | **8** | **0.10 (0.04 - 0.19)** | **13.67 (11.20 - 16.45)** |
| 0-2 months | 1243 |  | 320 | 25.74 (23.33 - 28.27) | 39 | 3.14 (2.24 - 4.26) | 3 | 0.24 (0.05 - 0.70) | 12.19 (8.81 - 16.28) |
| 3-5 months | 869 |  | 101 | 11.62 (9.57 - 13.94) | 18 | 2.07 (1.23 - 3.25) | 0 |  | 17.82 (10.92 - 26.70) |
| 6-8 months | 995 |  | 64 | 6.43 (4.99 - 8.14) | 11 | 1.11 (0.55 - 1.97) | 2 | 0.20 (0.02 - 0.72) | 17.19 (8.90 - 28.68) |
| 9-11 months | 1085 |  | 63 | 5.81 (4.49 - 7.37) | 6 | 0.55 (0.20 - 1.20) | 0 |  | 9.52 (3.58 - 19.59) |
| 12-17 months | 2042 |  | 82 | 4.02 (3.21 - 4.96) | 7 | 0.34 (0.14 - 0.71) | 2 | 0.10 (0.01 - 0.35) | 8.54 (3.50 - 16.80) |
| 18-23 months | 2024 |  | 65 | 3.21 (2.49 - 4.08) | 14 | 0.69 (0.38 - 1.16) | 1 | 0.05 (0.00 - 0.27) | 21.54 (12.31 - 33.49) |

| Total 0-17 years, full period (2) | 820404 | 2692 | 9611 | 1.17 (1.15 - 1.20) | 640 | 0.08 (0.07 - 0.08) | 84 | 0.01 (0.01 - 0.01) | 6.66 (6.17 - 7.18) |
| --- | --- | --- | --- | --- | --- | --- | --- | --- | --- |
| <1 year | 22518 | 1432 | 2952 | 13.11 (12.67 - 13.56) | 142 | 0.63 (0.53 - 0.74) | 15 | 0.07 (0.04 - 0.11) | 4.81 (4.07 - 5.65) |
| 1 year | 24797 | 1525 | 852 | 3.44 (3.21 - 3.67) | 44 | 0.18 (0.13 - 0.24) | 5 | 0.02 (0.01 - 0.05) | 5.16 (3.78 - 6.87) |
| 2 years | 23032 | 1384 | 471 | 2.04 (1.87 - 2.24) | 22 | 0.10 (0.06 - 0.14) | 4 | 0.02 (0.00 - 0.04) | 4.67 (2.95 - 6.99) |
| 3 years | 24573 | 1452 | 324 | 1.32 (1.18 - 1.47) | 14 | 0.06 (0.03 - 0.10) | 3 | 0.01 (0.00 - 0.04) | 4.32 (2.38 - 7.14) |
| 4 years | 24858 | 1484 | 265 | 1.07 (0.94 - 1.20) | 15 | 0.06 (0.03 - 0.10) | 6 | 0.02 (0.01 - 0.05) | 5.66 (3.20 - 9.16) |
| 5 years | 26494 | 1574 | 241 | 0.91 (0.80 - 1.03) | 18 | 0.07 (0.04 - 0.11) | 4 | 0.02 (0.00 - 0.04) | 7.47 (4.49 - 11.55) |
| 6 years | 31300 | 1881 | 225 | 0.72 (0.63 - 0.82) | 20 | 0.06 (0.04 - 0.10) | 1 | 0.00 (0.00 - 0.02) | 8.89 (5.51 - 13.39) |
| 7 years | 34857 | 2062 | 230 | 0.66 (0.58 - 0.75) | 14 | 0.04 (0.02 - 0.07) | 3 | 0.01 (0.00 - 0.03) | 6.09 (3.37 - 10.00) |
| 8 years | 39241 | 2325 | 253 | 0.64 (0.57 - 0.73) | 23 | 0.06 (0.04 - 0.09) | 3 | 0.01 (0.00 - 0.02) | 9.09 (5.85 - 13.33) |
| 9 years | 44703 | 2588 | 266 | 0.60 (0.53 - 0.67) | 24 | 0.05 (0.03 - 0.08) | 0 |  | 9.02 (5.87 - 13.13) |
| 10 years | 50126 | 2914 | 315 | 0.63 (0.56 - 0.70) | 31 | 0.06 (0.04 - 0.09) | 3 | 0.01 (0.00 - 0.02) | 9.84 (6.79 - 13.68) |
| 11 years | 54337 | 3120 | 307 | 0.56 (0.50 - 0.63) | 29 | 0.05 (0.04 - 0.08) | 4 | 0.01 (0.00 - 0.02) | 9.45 (6.42 - 13.28) |
| 12 years | 58210 | 3371 | 328 | 0.56 (0.50 - 0.63) | 26 | 0.04 (0.03 - 0.07) | 4 | 0.01 (0.00 - 0.02) | 7.93 (5.24 - 11.40) |
| 13 years | 61897 | 3621 | 374 | 0.60 (0.54 - 0.67) | 35 | 0.06 (0.04 - 0.08) | 6 | 0.01 (0.00 - 0.02) | 9.36 (6.61 - 12.77) |
| 14 years | 63855 | 3737 | 430 | 0.67 (0.61 - 0.74) | 49 | 0.08 (0.06 - 0.10) | 8 | 0.01 (0.01 - 0.02) | 11.40 (8.55 - 14.78) |
| 15 years | 70862 | 4091 | 477 | 0.67 (0.61 - 0.74) | 38 | 0.05 (0.04 - 0.07) | 7 | 0.01 (0.00 - 0.02) | 7.97 (5.70 - 10.77) |
| 16 years | 77353 | 4490 | 567 | 0.73 (0.67 - 0.80) | 40 | 0.05 (0.04 - 0.07) | 1 | 0.00 (0.00 - 0.01) | 7.05 (5.09 - 9.48) |
| 17 years | 87391 | 5043 | 734 | 0.84 (0.78 - 0.90) | 56 | 0.06 (0.05 - 0.08) | 7 | 0.01 (0.00 - 0.02) | 7.63 (5.81 - 9.79) |
| 1-4 years | 97260 | 1461 | 1912 | 1.97 (1.88 - 2.06) | 95 | 0.10 (0.08 - 0.12) | 18 | 0.02 (0.01 - 0.03) | 4.97 (4.04 - 6.04) |
| 5-11 years | 281058 | 2359 | 1837 | 0.65 (0.62 - 0.68) | 159 | 0.06 (0.05 - 0.07) | 18 | 0.01 (0.00 - 0.01) | 8.66 (7.41 - 10.04) |
| 12-17 years | 419568 | 4061 | 2910 | 0.69 (0.67 - 0.72) | 244 | 0.06 (0.05 - 0.07) | 33 | 0.01 (0.01 - 0.01) | 8.38 (7.40 - 9.45) |
| 18-49 years | 3339485 | 4702 | 86961 | 2.60 (2.59 - 2.62) | 10293 | 0.31 (0.30 - 0.31) | 2837 | 0.08 (0.08 - 0.09) | 11.84 (11.62 - 12.05) |
| 50-79 years | 2095032 | 3143 | 277377 | 13.24 (13.19 - 13.29) | 59911 | 2.86 (2.84 - 2.88) | 73672 | 3.52 (3.49 - 3.54) | 21.60 (21.45 - 21.75) |
| 80+ years | 349562 | 2948 | 141502 | 40.48 (40.32 - 40.64) | 16136 | 4.62 (4.55 - 4.69) | 116871 | 33.43 (33.28 - 33.59) | 11.40 (11.24 - 11.57) |

CI = confidence interval. 1. Analysis of a subset of cases with data on age in months under 2 years of age, weeks 2020-32 to 2021-39, from seven countries (Austria, Cyprus, Finland, Ireland, Luxembourg, Slovakia and Sweden). Case rates per 100 000 population are not provided for these age groups. 2. Full dataset for the period weeks 2020-32 to 2021-39. 3. Full dataset restricted to the period of Delta dominance (defined per country, median dates: 2021-28 to 2021-39).

Limiting to children aged 2-17 years, the overall risks of these outcomes fell to 0.8%, 0.06%, 7.5% and 0.01%, respectively. The risk of hospitalisation was highest among the youngest infants (0-2 months), with point estimates that decreased with increasing age in months or years to 9 years and then increased with each year from 12 to 17. Males aged 0-17 years were slightly more likely than females to be admitted to hospital (1.2% vs 1.1%, p<0.0001) or ICU (0.09% vs 0.07%, p<0.05), but these differences did not exist in all paediatric age groups. Results consistent with the full period were obtained for periods coinciding with delta dominance, although hospitalisation was more common among children <1 year (RISK 14.7% delta vs 13.1% full period, p<0.01; all other outcomes/age groups p>0.14).

### Stratification by presence of any comorbidity

Data on comorbidities were available for 210,008 out of 460,790 cases (45.6%) from seven countries. Among these, 203,548 (96.9%) reported having none, 5,773 (2.7%) one and 687 (0.3%) two or more comorbidities. Data on comorbidities were less likely to be reported for hospitalised than non-hospitalised paediatric cases (30.0% vs 45.8%, p<0.0001). Overall, crude risks for each outcome were higher among children with at least one comorbidity than none. After controlling for reporting country, period and sex, adjusted odds of hospitalisation, ICU admission and death were seven, nine and 27 times higher, respectively, among cases with at least one comorbidity compared to those with none (Table 2).

**Table 2:**
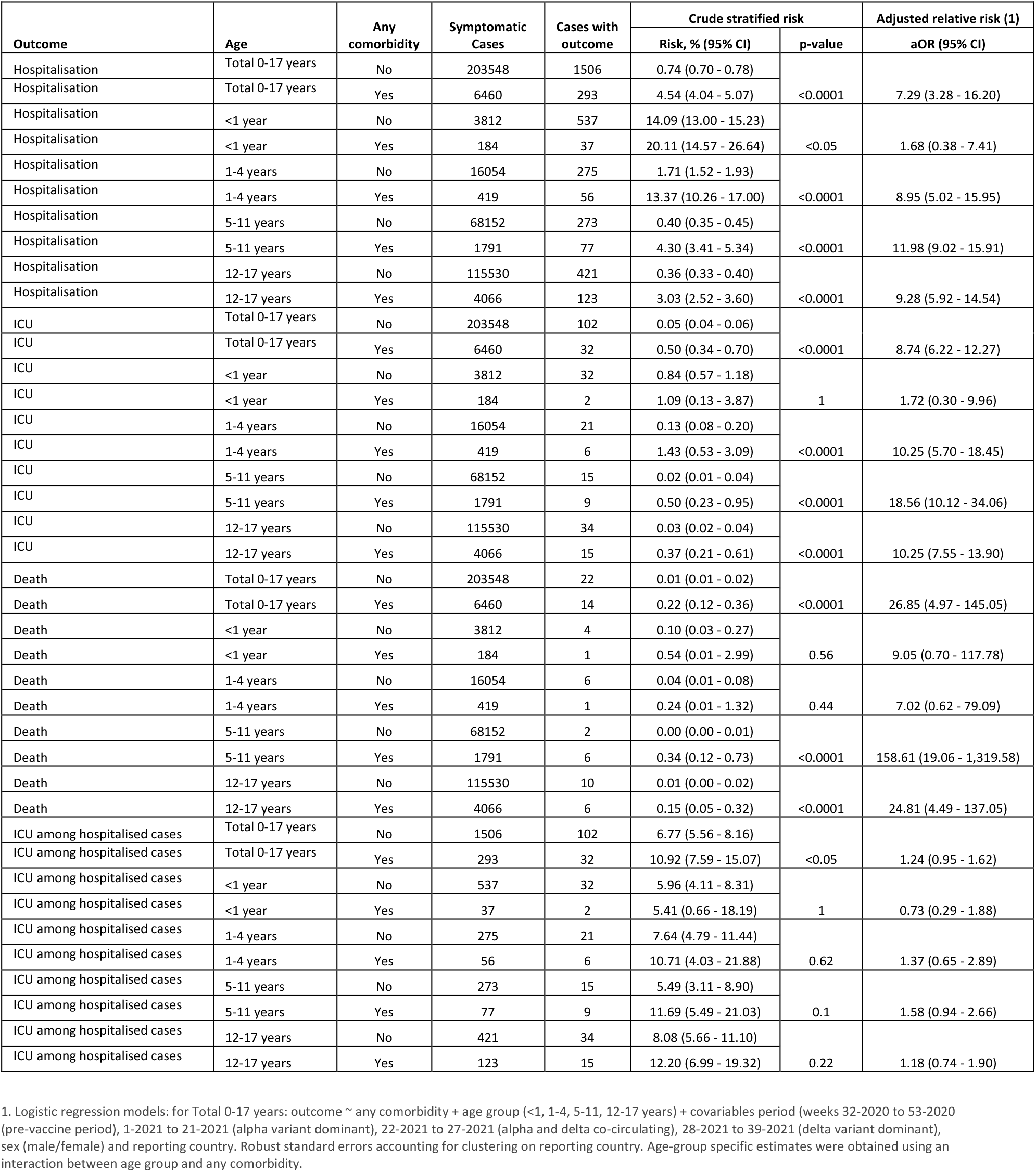
**Age-specific counts, crude risks and adjusted odds ratios for severe COVID-19 outcomes comparing cases with and without any comorbidity, pooled from seven EU countries, weeks 32-2020 to 39-2021**

| Outcome | Age | Any comorbidity | Symptomatic Cases | Cases with outcome | Crude stratified risk |  | Adjusted relative risk (1) |
| --- | --- | --- | --- | --- | --- | --- | --- |
|  |  |  |  |  | Risk, % (95% CI) | p-value | aOR (95% CI) |
| Hospitalisation | Total 0-17 years | No | 203548 | 1506 | 0.74 (0.70 - 0.78) | <0.0001 | 7.29 (3.28 - 16.20) |
| Hospitalisation | Total 0-17 years | Yes | 6460 | 293 | 4.54 (4.04 - 5.07) |  |  |
| Hospitalisation | <1 year | No | 3812 | 537 | 14.09 (13.00 - 15.23) | <0.05 | 1.68 (0.38 - 7.41) |
| Hospitalisation | <1 year | Yes | 184 | 37 | 20.11 (14.57 - 26.64) |  |  |
| Hospitalisation | 1-4 years | No | 16054 | 275 | 1.71 (1.52 - 1.93) | <0.0001 | 8.95 (5.02 - 15.95) |
| Hospitalisation | 1-4 years | Yes | 419 | 56 | 13.37 (10.26 - 17.00) |  |  |
| Hospitalisation | 5-11 years | No | 68152 | 273 | 0.40 (0.35 - 0.45) | <0.0001 | 11.98 (9.02 - 15.91) |
| Hospitalisation | 5-11 years | Yes | 1791 | 77 | 4.30 (3.41 - 5.34) |  |  |
| Hospitalisation | 12-17 years | No | 115530 | 421 | 0.36 (0.33 - 0.40) | <0.0001 | 9.28 (5.92 - 14.54) |
| Hospitalisation | 12-17 years | Yes | 4066 | 123 | 3.03 (2.52 - 3.60) |  |  |
| ICU | Total 0-17 years | No | 203548 | 102 | 0.05 (0.04 - 0.06) | <0.0001 | 8.74 (6.22 - 12.27) |
| ICU | Total 0-17 years | Yes | 6460 | 32 | 0.50 (0.34 - 0.70) |  |  |
| ICU | <1 year | No | 3812 | 32 | 0.84 (0.57 - 1.18) | 1 | 1.72 (0.30 - 9.96) |
| ICU | <1 year | Yes | 184 | 2 | 1.09 (0.13 - 3.87) |  |  |
| ICU | 1-4 years | No | 16054 | 21 | 0.13 (0.08 - 0.20) | <0.0001 | 10.25 (5.70 - 18.45) |
| ICU | 1-4 years | Yes | 419 | 6 | 1.43 (0.53 - 3.09) |  |  |
| ICU | 5-11 years | No | 68152 | 15 | 0.02 (0.01 - 0.04) | <0.0001 | 18.56 (10.12 - 34.06) |
| ICU | 5-11 years | Yes | 1791 | 9 | 0.50 (0.23 - 0.95) |  |  |
| ICU | 12-17 years | No | 115530 | 34 | 0.03 (0.02 - 0.04) | <0.0001 | 10.25 (7.55 - 13.90) |
| ICU | 12-17 years | Yes | 4066 | 15 | 0.37 (0.21 - 0.61) |  |  |
| Death | Total 0-17 years | No | 203548 | 22 | 0.01 (0.01 - 0.02) | <0.0001 | 26.85 (4.97 - 145.05) |
| Death | Total 0-17 years | Yes | 6460 | 14 | 0.22 (0.12 - 0.36) |  |  |
| Death | <1 year | No | 3812 | 4 | 0.10 (0.03 - 0.27) | 0.56 | 9.05 (0.70 - 117.78) |
| Death | <1 year | Yes | 184 | 1 | 0.54 (0.01 - 2.99) |  |  |
| Death | 1-4 years | No | 16054 | 6 | 0.04 (0.01 - 0.08) | 0.44 | 7.02 (0.62 - 79.09) |
| Death | 1-4 years | Yes | 419 | 1 | 0.24 (0.01 - 1.32) |  |  |
| Death | 5-11 years | No | 68152 | 2 | 0.00 (0.00 - 0.01) | <0.0001 | 158.61 (19.06 - 1,319.58) |
| Death | 5-11 years | Yes | 1791 | 6 | 0.34 (0.12 - 0.73) |  |  |
| Death | 12-17 years | No | 115530 | 10 | 0.01 (0.00 - 0.02) | <0.0001 | 24.81 (4.49 - 137.05) |
| Death | 12-17 years | Yes | 4066 | 6 | 0.15 (0.05 - 0.32) |  |  |
| ICU among hospitalised cases | Total 0-17 years | No | 1506 | 102 | 6.77 (5.56 - 8.16) | <0.05 | 1.24 (0.95 - 1.62) |
| ICU among hospitalised cases | Total 0-17 years | Yes | 293 | 32 | 10.92 (7.59 - 15.07) |  |  |
| ICU among hospitalised cases | <1 year | No | 537 | 32 | 5.96 (4.11 - 8.31) | 1 | 0.73 (0.29 - 1.88) |
| ICU among hospitalised cases | <1 year | Yes | 37 | 2 | 5.41 (0.66 - 18.19) |  |  |
| ICU among hospitalised cases | 1-4 years | No | 275 | 21 | 7.64 (4.79 - 11.44) | 0.62 | 1.37 (0.65 - 2.89) |
| ICU among hospitalised cases | 1-4 years | Yes | 56 | 6 | 10.71 (4.03 - 21.88) |  |  |
| ICU among hospitalised cases | 5-11 years | No | 273 | 15 | 5.49 (3.11 - 8.90) | 0.1 | 1.58 (0.94 - 2.66) |
| ICU among hospitalised cases | 5-11 years | Yes | 77 | 9 | 11.69 (5.49 - 21.03) |  |  |
| ICU among hospitalised cases | 12-17 years | No | 421 | 34 | 8.08 (5.66 - 11.10) | 0.22 | 1.18 (0.74 - 1.90) |
| ICU among hospitalised cases | 12-17 years | Yes | 123 | 15 | 12.20 (6.99 - 19.32) |  |  |

1. Logistic regression models: for Total 0-17 years: outcome ~ any comorbidity + age group (<1, 1-4, 5-11, 12-17 years) + covariables period (weeks 32-2020 to 53-2020 (pre-vaccine period), 1-2021 to 21-2021 (alpha variant dominant), 22-2021 to 27-2021 (alpha and delta co-circulating), 28-2021 to 39-2021 (delta variant dominant), sex (male/female) and reporting country. Robust standard errors accounting for clustering on reporting country. Age-group specific estimates were obtained using an interaction between age group and any comorbidity.

### Distribution of comorbidities among hospitalised cases

Of the 210,008 paediatric cases with data on comorbidities, 83.7% (1,506/1,799) of those hospitalised were reported not to have any. This proportion fell with increasing age: 93.6% (<1 year), 83.1% (1-4 years), 78.0% (5-11 years) and 77.4% (12-17 years). Among 293 (16.3%) hospitalised cases reporting a total of 314 comorbidities (19 cases had more than one), the most frequently reported conditions were cancer (20.7%), diabetes (18.2%), cardiac (15.9%) and lung (11.8%) disease (Figure 2b). Cancer (n=7) followed by HIV (5), lung disease (5), cardiac disease (4) and diabetes (4) were most common among the 34/136 (25.0%) cases admitted to ICU reporting at least one comorbidity, and 14/36 fatal cases had at least one comorbidity.

**Figure 2:**
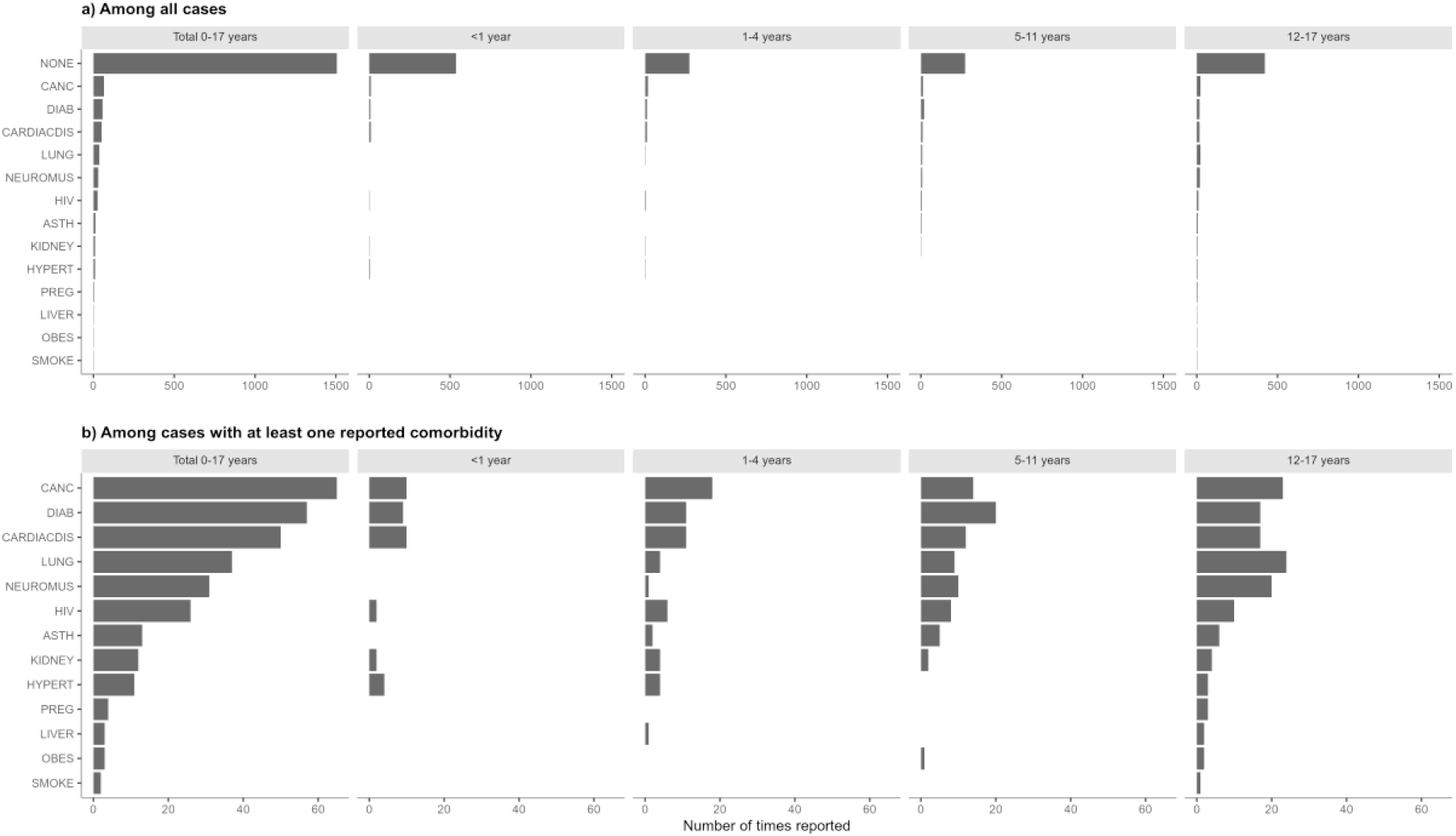
Occurrence of reported comorbidities among hospitalised paediatric COVID-19 cases by age group, pooled from seven EU countries, weeks 32-2020 to 39-2021, a) among all cases and b) among cases with at least one reported comorbidity. Notes: CANC=Cancer; DIAB=Diabetes; CARDIACDIS=Cardiac disease; LUNG=Lung disease; NEUROMUS=neuromuscular disease; HIV=human immunodeficiency virus; ASTH=asthma; KIDNEY=kidney disease; HYPERT=hypertension; PREG=pregnancy (including post-partum); LIVER=liver disease; OBES=obesity; SMOKE=smoker/history of smoking. One instance each of smoker and pregnancy reported in two infants aged 0 months (<1 year) are not shown in the figure since it was not possible to confirm if this coding related to the mother.

## Discussion

As of week 47-2021 only 15.2% (range: 1.0-29.0%) of children <18 years of age in the EU/EEA (22.1%; range 7.2-25.5% in the 10 countries included in this study) have been fully vaccinated against COVID-19 [5]. The European Medicines Agency (EMA) is currently evaluating the emergency authorisation of Comirnaty in children aged 5-11 years old [6]. The evidence presented here on the burden of COVID-19 in children indicates that case notification and hospital admission rates among children rise as transmission increases, but that most children with symptomatic COVID-19 have a very low risk of death or hospitalisation. For every 10,000 symptomatic paediatric cases reported during the, approximately 117 were hospitalised and eight required ICU admission or respiratory support.

The high incidence in many parts of the EU/EEA mean large numbers of unvaccinated children are likely to be exposed to the virus, leading to increases in the absolute numbers of children with severe COVID-19 outcomes. In addition, perhaps particularly for countries that have achieved high levels of vaccination coverage in adults, the bulk of community transmission could be increasingly among children [7]. Thus, tailored national guidance and appropriate mitigation measures, notably in schools and other places where children congregate, will continue to be essential [1].

This study highlights that while the individual risk of a severe COVID-19 outcome is low among healthy children, these risks are substantially elevated for children with underlying risk factors [8]. Despite the low individual risks to healthy children, 83.7% of all hospitalised childhood COVID-19 cases had no reported comorbidity, demonstrating the additive impact of high levels of community transmission.

The elevated risks of hospitalisation observed for children less than 2 years of age likely reflects low threshold for admission in this age group, however the increased risk and data on the presence of SARS-CoV-2 antibodies in breastmilk need to be studied further urgently to support paediatric clinical decision-making and the provision of information on vaccination to pregnant women [9]. Further research is required also to clarify risk factors for severe outcomes among children, including <2 years of age.

This analysis did not consider other health impacts of COVID-19 on children such as the prevalence and burden of paediatric inflammatory multisystem syndrome (PIMS) or post-COVID-19 syndrome in children, as well as the numerous indirect negative health and mental health impacts on children caused by disruptions to their social and educational lives [1]. Such factors would, nonetheless, be important considerations for decision-making about vaccination of children [10].

There are important limitations to this study. The analysis is based on surveillance data reported to TESSy. Data on vaccination status of the children in this study was not available, however vaccination rates in children were very low generally and vaccines only approved for use in children 12-17 years at the time of the study. As children are less likely to be symptomatic for COVID-19 than adults [11,12], reporting of cases may be biased towards those with severe disease, and although cases with hospitalisation date prior to date of onset were considered not hospitalised for this analysis, some remaining reported paediatric hospitalisations may have been for reasons other than COVID-19. While this could over-estimate the crude risk of hospitalisation, we are aware that hospitalisations are under-reported to TESSy by at least three of the countries in our study, which would have the opposite effect, and our results are comparable to those reported among symptomatic cases in a recent review [10]. Reporting of comorbidities was less likely among cases with severe outcomes, making our effect estimates conservative for cases with a comorbidity. Low numbers of children with severe outcomes diminish the analytical power of the study, particularly for less common outcomes. This, together with incomplete reporting of comorbidities, prevented a detailed risk factor analysis.

## Conclusions

COVID-19 hospital admissions in children increase in line with increasing case numbers. Individual risks of a severe COVID-19 outcome are substantially elevated for those with a comorbidity compared to healthy children, however most children hospitalised in this study had no reported comorbidity. This demonstrates the additive impact of high levels of community transmission and can inform decision-making around which groups of children to offer COVID-19 vaccination.

Preventive measures to reduce transmission and severe outcomes in children remain critical.

## Data Availability

All data produced in the present study are available upon reasonable request to the authors

## Declaration of interest

None to report

## Acknowledgements

We thank the TESSy data managers for their ongoing support and all people involved in the ECDC COVID-19 response. Special thanks are due to ECDC staff who work tirelessly on the weekly processing and analysis of TESSy data, who have provided input to or reviewed this work, in particular Tommi Karki, Enrique Delgado, Gaetano Marrone, Pasi Penttinen and Ole Heuer. We would like to acknowledge Richard Pebody, Piers Mook and Maarten Vanhaverbeke from the WHO Regional Office for Europe for their ideas and critical review which help shaped this work. The European COVID-19 surveillance network is jointly coordinated by the ECDC and the WHO Regional Office for Europe. The authors are grateful to all network members and National Focal Points for Viral Respiratory Disease for kindly collecting, uploading and helping in the interpretation of weekly European surveillance data. We particularly thank experts from the countries included in this study (listed in alphabetical order): Austria (Stephan Aberle, Monika Redlberger-Fritz and Daniela Schmid), Cyprus (Costas Constantinou, Ioanna Gregoriou, Christos Karagiannis, and Barbara Zinieri), Finland (Idil Hussein, Niina Ikonen, Maia Jeganova, Jeremia Kapanen, Mia Kontio, Jan-Erik Löflund, Merit Melin and Carita Savolainen-Kopra), Germany (Doris Altmann, Christian Drosten, Andreas Tille), Ireland (Jeff Connell, John Cuddihy, Gillian Cullen, Lorraine Doherty, Lisa Domegan, Linda Dunford, Margaret Fitzgerald, Patricia Garvey, Derval Igoe, Sarah Jackson, Jolita Mereckiene, Niamh Murphy and Joan O’Donnell), Italy (Angela Di Martino, Simona Puzelli, Flavia Riccardo, Caterina Rizzo and Paola Stefanelli), Luxembourg (Tamir Abdelrahman, Joël Mossong and Gerard Scheiden), Malta (Christopher Barbara, Maria Louise Borg, Warren Bruno, Charmaine Gauci, Jackie Maistre Melillo and Tanya Melillo), Slovakia (Ivan Bakoss, Eva Chmelanova, Ivan Kapitáň, Ján Mikas, Jana Námešná, Jozef Nováček, Maria Ondekova and Edita Staronová) and Sweden (Sören Andersson, Mia Brytting, AnnaSara Carnahan, Shaman Muradrasoli, Moa Rehn and Katherina Zakikhany).

## Author contributions

NB, ND, CD, AP, JS conceived the study. NB conducted the data analysis. The country study group authors conducted COVID-19 surveillance and data collections in their respective countries. NB, CD, AP, JS drafted the manuscript, with all authors providing input and contributing to its finalisation.

## Data sharing statement

All relevant data are within the paper. Access to data requests for subsets of COVID-19 TESSy datasets can be requested from the European Centre for Disease Prevention and Control.

## Notes

### Competing Interest Statement

The authors have declared no competing interest.

### Funding Statement

This study did not receive any funding

### Author Declarations

The analysis is based on surveillance data reported to the European Surveillance System (TESSy) (www.ecdc.europa.eu and https://www.ecdc.europa.eu/en/covid-19/country-overviews)

